# The mental health of staff working in intensive care during COVID-19

**DOI:** 10.1101/2020.11.03.20208322

**Authors:** Neil Greenberg, Dale Weston, Charlotte Hall, Tristan Caulfield, Victoria Williamson, Kevin Fong

## Abstract

**Background:** Intensive Care Unit (ICU), anaesthetic and theatres staff have faced significant challenges during the COVID-19 pandemic which have the potential to adversely affect their mental health

**Aims:** To identify the rates of probable mental health disorder in ICU and anaesthetic staff in six English hospitals during June and July 2020

**Methods:** An anonymised brief web-based survey comprising standardised questionnaires examining depression, anxiety symptoms, symptoms of Post Traumatic Stress Disorder (PTSD), wellbeing and alcohol use was administered to staff.

**Results:** 709 participants completed the surveys comprising 291 (41%) doctors, 344 (48.5%) nurses, and 74 (10.4%) other healthcare staff. Over half (58.8%) reported good wellbeing, however 45.4% met the threshold for probable clinical significance on at least one of the following measures: severe depression (6.3%), PTSD (39.5%), severe anxiety (11.3%) or problem drinking (7.2%). 13.4% of respondents reported frequent thoughts of being better off dead, or of hurting themselves in the past two weeks. We found that doctors consistently reported better mental health than nurses.

**Conclusions:** We found substantial rates of probable mental health disorders, and thoughts of self-harm, amongst ICU staff; these difficulties were especially prevalent in nurses. These results indicate the need for a national strategy to protect the mental health, and decrease the risk of functional impairment, of ICU staff whilst they carry out their essential work during COVID-19.

**Occ Med Statements: article should contain 3 statements (each max of 3 bullets of max 50 words each):** *What is already known about this subject:*

- Intensive care unit (ICU) staff are regularly exposed to traumatic situations as part of their job
- Previous studies have shown them to be at risk of psychological and moral distress
- Little is known about the mental health of ICU staff during the current pandemic What this study adds

- Almost half of ICU staff report symptoms consistent with a probable diagnosis of post traumatic stress disorder, severe depression or anxiety or problem drinking
- Around 1 in 7 ICU staff report recent thoughts of self-harm or of wanting to be better off dead
- Nursing staff are more likely to report higher levels of distress than doctors or other clinical staff

What impact this may have on practice or policy

- Healthcare managers need to prioritise staff mental health support and timely access to evidence based treatments for ICU staff
- Supervisors and managers should be aware that a substantial proportion of ICU staff may perform less well because of their current poor state of mental health
- More work is needed to understand whether the high levels of mental health symptoms identified in this study are truly indicative of high levels of clinical need for mental healthcare

## Introduction

The COVID-19 virus outbreak was declared a pandemic on March 12, 2020 by the World Health Organisation [1]. Across the globe healthcare workers have been at the frontline of each nation’s response, labouring to meet a sudden and dramatic increase in demand and workload across the full spectrum of healthcare. Among those most directly impacted have been intensive care and anaesthetic teams who together augmented and expanded critical care provision.

Frontline healthcare staff experience myriad psychological stressors, including fears of contracting the virus and endangering their loved ones, concerns over the lack of personal protective equipment (PPE), and distress relating to adverse patient outcomes and loss of patient lives despite their best efforts [2,3].

Within the UK a substantial proportion of the 175,000 patients admitted to hospital with COVID-19 are admitted for critical care in acute hospitals. To accommodate this unprecedented surge, during the first wave of the pandemic in the UK, hospitals were forced to create ad hoc intensive care units (ICUs) with heavily modified staffing models; reducing the usual 1:1 ICU nurse patient ratio to as low as 1:6 in some cases [4]. Pre-existing shortages of experienced ICU staff have been greatly exacerbated by high levels of staff sickness and quarantine during the first COVID-19 surge.

Consequently, ICU staff have faced a particularly challenging time frequently working in areas where the risk of COVID-19 exposure is high for long periods, wearing PPE, with the challenges of managing staff and equipment shortages on a daily basis especially during the first wave. At times this had made it difficult for staff to deliver a normal standards of care. The high rate of mortality amongst COVID-19 patients admitted to ICU, coupled with difficulty in communication and providing adequate end-of-life support to patients and their next of kin, because of visiting restrictions, has been a specific stressor for all staff working in ICU.

These working conditions have the potential to adversely impact the mental health of ICU staff, including the experience of psychological distress, moral injury, [5] and the development of mental health difficulties such as depression and post-traumatic stress disorder (PTSD). In order to ascertain what level of psychological support may be necessary, an survey of the mental health of frontline staff working in ICU settings during COVID-19 in June and July 2020.

## Methods

Intensive care units, across six NHS hospitals with peak ICU bed occupancy figures ranging between 10 and 75 critically ill COVID-19 patients, were identified from ICNARC (Intensive Care National Audit and Research Centre) data and local ICU reporting systems. The hospitals were drawn from a range of NHS acute trusts including two metropolitan teaching hospitals and four district general hospitals. The data were gathered as part of a service evaluation exercise, in an attempt to monitor the effect of heavily modified working patterns on intensive care and anaesthetic staff during the UK’s first COVID-19 surge.

We engaged with clinical leads from participating ICUs and encouraged the circulation and completion of the online survey. The survey was distributed via departmental email mailing lists and cascaded through departmental SMS contact groups.

A brief online survey tool – designed to be completed in less than 5 minutes - was compiled, comprising a number of validated questions assessing the mental health status and psychological well-being.

The survey comprised the following measures, for which binary outcomes variables were defined using the following cut-off scores to indicate a case; the 7-item Generalised Anxiety Disorder (GAD) scale to measure probable moderate anxiety disorder with a score >9 indicating a probable moderate anxiety disorder and >15 indicating probable severe anxiety disorder [6]; the 9-item Patient Health Questionnaire (PHQ-9) with a score of >9 indicating probable moderate depression and >19 probable severe depression [7]; the 6-item Post-Traumatic Stress Disorder checklist (PCL-6) civilian version to measure PTSD [8] with a score of >13 indicating the presence of probable PTSD and the AUDIT-C with a score of >7 indicating problem drinking [9]. We also examined participants’ responses as to whether they had had “thoughts that [they] would be better off dead, or of hurting [themselves] in some way [in the past two weeks]” which is a single item within the PHQ9 questionnaire. Finally the Warwick Edinburgh Mental Wellbeing Scale (WEMWBS) [10], a 14 item scale where all items are worded positively, was used to explore feelings and functioning aspects of mental wellbeing. These questionnaires were incorporated into an online survey form, which could be accessed from a hyperlink embedded within an email or SMS message.

The brief online survey was anonymous at the point of collection and the resultant data was uncoupled from identifying detail from the originating device. Participants completed the survey voluntarily, with the knowledge that the data would be anonymised, and were free to stop at any point during their completion of the survey. Incomplete surveys were discarded.

The survey was built using the LimeSurvey tool (https://www.limesurvey.org/) and hosted on a dedicated secure university server. No registration was needed to participate in the survey and no individually-identifying details were collected from participants.

The need for ethical review was discussed with two university ethics committees both of which confirmed that, as an anonymised audit and quality improvement exericise, the survey did not require ethical approval. The ethical committees note that participants were not randomised, the evaluation protocol did not demand any change in care provision or any particular intervention, and the findings would not be generalisable outside of the staff group that were the focus of the survey. The NHS Health Research Authority ‘is my study research?’ decision tool also confirmed that the study did not require review by a research ethics committee.

Descriptive analyses were conducted to provide an overview of the sample characteristics. Bivariate correlations were used to examine the relationship between mental health measure scores. We examined differences in scores across the different professions (doctors, nurses, and other healthcare workers in ICU) using logistic regression analyses.

## Results

Overall, 709 participants took part in the study. Of these, 291 (41%) identified themselves as being doctors, 344 (48.5%) nurses, and 74 (10.4%) as being in other clinical roles.

The majority of participants reported good wellbeing on the WEMWBS (n= 418, 58.8%), although almost half of participants (*n* = 322, 45.4%) met the threshold for at least one of the following measures: severe depression, PTSD, severe anxiety or problem drinking (see Table 1).

**Table 1:** Frequencies of participants split by role which met psychological measures thresholds; Logistic regressions carried out on each psychological measure threshold to examine effect of role are presented.

|  | N<br>(% of<br>sample) | Role |  |  | Logistic Regression |  |  |
| --- | --- | --- | --- | --- | --- | --- | --- |
|  |  | Counts |  |  | 95% CI for Odds Ratio |  |  |
|  |  | Doctor<br>(% of<br>sample) | Nurse<br>(% of<br>sample) | Other<br>(% of<br>sample) | B (SE) | Lower | Odds Ratio |
| Good Wellbeing | 417 (58.8) | 185 (63.6) | 186 (54.1) | 46 (62.16) | .39*<br>(.16) | 1.08 | 1.48 |
| Moderate Depression | 262 (36.9) | 76 (26.1) | 167 (48.5) | 19 (25.7) | -0.98*<br>(0.17) | 0.27 | 0.38 |
| Probable PTSD | 280 (39.5) | 92 (31.6) | 168 (48.8) | 20 (27.0) | -0.73*<br>(0.16) | 0.35 | 0.48 |
| Severe Depression | 45 (6.3) | 13 (4.5) | 30 (8.7) | 2 (2.7) | -0.71*<br>(0.34) | 0.25 | 0.49 |
| Moderate Anxiety | 189 (26.7) | 58 (19.9) | 115 (33.4) | 16 (21.6) | -0.70*<br>(0.19) | 0.34 | 0.49 |
| Severe Anxiety | 80 (11.3) | 23 (7.9) | 52 (15.1) | 5 (6.8) | -0.73*<br>(.26) | 0.29 | 0.48 |
| Problem Drinking | 51 (7.2) | 20 (6.9) | 28 (8.1) | 3 (4.1) | -.18<br>(.30) | 0.46 | 0.83 |
| AMD | 322 (45.4%) |  |  |  |  |  |  |

Note. AMD = Any Mental Disorder (consisting of at least one of the following: Severe Depression, Severe Anxiety, Probable PTSD or Problem Drinking. Good Wellbeing is indicative of a score of $\geq 43$ on WEMWBS; Moderate Depression equates to a score of $\geq 10$ ; and Severe Depression equates to a score of $\geq 20$ on the PHQ9; Probable PTSD equates to a score of $\geq 15$ on PCL6; Moderate Anxiety equates to a score of $\geq 11$ ; and Severe Depression equates to a score of $\geq 16$ on the GAD7; Problem drinking equates to a score of $\geq 8$ on AUDITC. The negative beta coefficient and odds ratio of less than 1 is due to coding of the Role predictor with Nurses as the reference category (thus Doctors = 1, Nurses = 0).

13.4% of respondents reported having thoughts that [they] would be better off dead, or of hurting [themselves] in some way several days or more frequently in the past two weeks. When examined by role a significantly higher proportion of nurses (19.2%) than doctors (7.6%) or other clinical staff (9.5%) (χ^2^ = 26.8, d.f.8, *p* < 0.002) reported these thoughts.

Logistic regression indicated that doctors were more likely, than other clinicians, to report good wellbeing and nurses were more likely to meet the thresholds for depression (moderate and severe), probable PTSD, and anxiety (moderate and severe) (see Table 1).

Higher scores on the WEMWBS were significantly associated with lower scores on all the other outcomes measures (depression, PTSD, anxiety and alcohol use). Measures of anxiety, depression and PTSD symptoms were significantly correlated with each other. No significant associations were found between any measure of poor mental health and alcohol consumption (see Table 2).

**Table 2:**
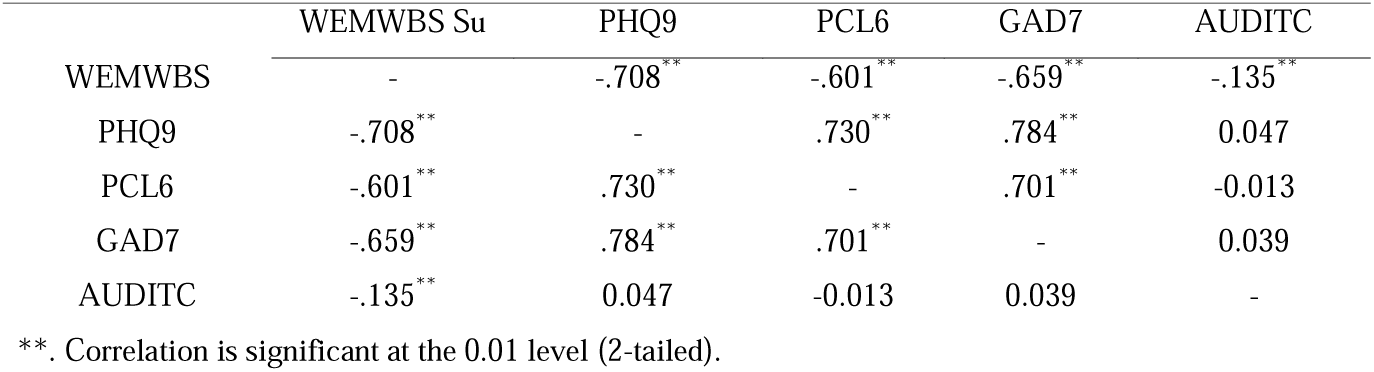
Bivariate correlations carried out between psychological measures.

## Discussion

We examined the mental health of impact of working in ICU settings during COVID-19 pandemic surge for NHS staff during June and July 2020. We identified a number of key finding most notably high rates of probable mental ill health with around 45% of the sample self-reporting symptoms of probable PTSD, severe depression or a severe anxiety disorder. More than 1 in 7 ICU staff reported thoughts that they would be better off dead, or of hurting themselves in some way several days over the past two weeks with nurses being more likely to report poor mental health and ideas of self harm or suicidal ideation than doctors or other healthcare staff. Lastly, although around 8% of the sample appeared to be at risk of alcohol related difficulties, this level of drinking was not significantly associated with poorer mental health outcomes.

Our results highlight the potential profound impact that COVID-19 has had on the mental health of frontline UK staff. The 2014 Adult Psychiatric Morbidity Study [11] found rates of probable PTSD in the UK general public to be approximately 4.4% and other studies have reported an overall PTSD prevalence in UK military personnel of around 6.5% with the highest rate, of 17%, in veterans who had recently served in a combat role [12]. Thus, probable PTSD rate we report (39.5%) was around nine times that found within the general population and more than double that found in recent combat veterans. Whilst further validation studies are required to better understand what proportion would actually meet diagnostic criteria for PTSD on clinical assessment, these data suggest that ICU clinicians are at a significantly elevated risk of suffering with PTSD. Our findings of high levels of PTSD, and other mental health difficulties such as depressive anxiety disorders, are highly relevant given that there is strong evidence poor mental health is associated with functional impairment which would increase the risk of patient safety incidents [13].

Whilst it is not possible to be certain why ICU clinicians reported such high levels of poor mental health, during the time these data were collected (June-July 2020), staff still faced a number of substantial stressors including long shifts, caring for dependent children and other household responsibilities [14] and regular exposure to ethical dilemmas with the consequential risk of moral injury [15]; some may also have still experienced difficulties with a lack of personal protective equipment (PPE) [13]. Whilst it is possible that the high levels of probable mental disorders are a result of ICU having always been a challenging environment, a 2015 study of 335 ICU staff found rates of probable PTSD of 8% amongst staff working with adults and 17% amongst staff working with children [16] suggesting the rates in this study are indeed elevated.

We found nurses were more likely to report experiencing mental health difficulties than doctors or other ICU staff. Whether this occupational group is more vulnerable to mental ill health by virtue of demographic risk factors, or whether other factors are unduly affecting this group, remains unclear. However, we note that UK ICU nurses are more likely to be younger adults and female [16] and this demographic has been shown to be at increased risk of suffering with poor mental health during the pandemic [17]. Other recent reports have also highlighted nurses as being at considerable risk of burnout and that nurses were at risk of suffering with poor mental health that was likely to affect retention rates [18].

Our finding that more than 1 in 7 clinicians (and nearly 1 in 5 nurses) working in ICU reported thoughts of self harm or suicide is also highly concerning. However, our survey did not ask whether respondents had made plans to carry out self-injurious or suicidal behaviours. It is also unclear how common such thoughts might be in people who become healthcare workers. For instance, a 2014 paper reported that around 14% of nursing students reported thoughts that made them a substantial suicide risk [19]. Whatever, the cause of such thoughts, we suggest that it is important that healthcare managers are aware of them and that measures to compassionately support any staff member at risk of suicide are put in place in a timely manner.

Perhaps unsurprisingly, our data showed that people who met the threshold for one form of probable mental disorder were substantially more likely to meet the threshold for another disorder. The exception to this was for alcohol misuse. Whilst, we identified around 10% of the sample reported drinking in a manner consistent with alcohol misuse, we found no association between poor mental health and alcohol misuse suggesting that within this sample self-medication with alcohol was not common. Also, the identified rate of alcohol misuse was very much in keeping with previous estimates of in healthcare staff which range from 1.6% to 24% [21]. This finding may have been because already tired and distressed staff recognised that excessive alcohol consumption would have made their ability to cope at work the next day even more difficult or because as healthcare staff they recognised the dangers of excessive alcohol use. Whatever the reason, this finding is heartening.

This study has several strengths and limitations. Amongst the strengths are the inclusion of a number of hospitals across the UK and completion of study assessments anonymously. A weakness of this study is the lack of participant demographic details which was done both for brevity and to preserve anonymity. As females, younger adults and those with dependent children are more likely to experience psychological difficulties, this information would be valuable in future investigations. Second, this study used self-report measures of mental illness rather than the gold-standard diagnostic interviews. Finally, it is possible that response bias occurred and those who participated had especially salient mental health difficulties they wanted to report. Future studies would be improved if either participants were randomly selected or a non-repsonder analysis was carried out.

Despite these limitations, the results of this study allow for several recommendations. First, our results suggest that NHS managers should prioritise provision of evidence based staff support which is likely both to improve psychological wellbeing and decrease the likelihood of psychologically unwell staff delivering substandard care. Second, it is also necessary to ensure that rapid access to formal treatment is available given its long term positive benefits (e.g. reduced staff absence, improved quality of life). Third, supervisor and peer support has been found to be particularly beneficial in supporting other trauma-exposed occupations such as for firefighters [22], or military personnel. Ensuring that a proportion of ICU staff receive active listen skills, or peer support training, may thus also be beneficial. Fourth, NHS managers should actively monitor the wellbeing of ICU staff in order that the impact of workload changes are properly understood and mitigated where possible. This would allow for staffing and other support measures to be implemented in a dynamic fashion ensuring the provision of high quality care whilst protecting the mental health of ICU staff upon whom the UK response to the pandemic depends.

This study identified that staff working in ICU during the current pandemic report substantially raised levels of poor mental health, in particular high rates of probable PTSD. The increased risk was particularly evident amongst nursing staff. Given the requirement for ICU staff to highly functional as they care for critically ill pateints, these data sugges that is imperative to ensure that adequate support is provisioned by NHS employers who have a moral and legal duty to appropriately safeguard staff wellbeing. Furthermore, unless employers properly protect the mental health of ICU staff, then they are more likely to function poorly with a consequential impact on their ability to deliver high quality patient care which is needed now more than ever.

### Contributions to the work

NG and KF designed the study and compiled the survey tool used. TC built the survey tool and extracted and coded the data provided by participants. NG, DW and CH analysed the data and built the results section which was also contributed to and interpreted by VW and KF. VW led on the introduction and discussion sections of the paper and all authors contributed to the methods section and edited the final manuscript and agreed to the final draft being submitted and agree to be accountable for all aspects of the work in ensuring that questions related to the accuracy or integrity of any part of the work are appropriately investigated and resolved.

## Data Availability

Data is not available for sharing.

